# Genetic and neurodevelopmental markers in schizophrenia-spectrum disorders: analysis of the combined role of the Cannabinoid Receptor 1 gene (*CNR1*) and dermatoglyphics

**DOI:** 10.1101/2024.01.23.24301648

**Authors:** Maria Guardiola-Ripoll, Alejandro Sotero-Moreno, Boris Chaumette, Oussama Kebir, Noemí Hostalet, Carmen Almodóvar-Payá, Mónica Moreira, Maria Giralt-López, Marie Odile-Krebs, Mar Fatjó-Vilas

**Author notes:** Co-last authors (Marie Odile-Krebs, Mar Fatjó-Vilas).

## Abstract

The aetiology of schizophrenia-spectrum disorders (SSD) involves genetic and environmental factors impacting neurodevelopmental trajectories. Dermatoglyphic pattern deviances have been associated with SSD and considered vulnerability markers for these disorders based on the shared ectodermal origin of the epidermis and the central nervous system. The endocannabinoid system participates in epidermal differentiation, is sensitive to the prenatal environment and is associated with SSD. We assessed whether the *Cannabinoid receptor 1* (*CNR1*) gene is a common denominator in dermatoglyphic pattern configurations and SSD risk and whether it modulates the dermatoglyphics-SSD association.

In a sample of 112 controls and 97 SSD patients, three dermatoglyphic markers were assessed: the total palmar a-b ridge count (TABRC), the a-b ridge count fluctuating asymmetry (ABRC-FA), and the pattern intensity index (PII). Two *CNR1* polymorphisms were genotyped: rs2023239-A/G and rs806379-A/T. We tested the *CNR1* association with SSD and with the dermatoglyphic variability within diagnostic groups. Secondly, we assessed the *CNR1* x dermatoglyphic measures interaction on SSD susceptibility.

Both polymorphisms were associated with the risk for SSD, and within controls, rs2023239 and rs806379 modulated the PII and TABRC, respectively. Lastly, our data showed that rs2023239 modulated the relationship between PII and SSD: a high PII score was associated with a lower SSD risk within G-allele-carriers and a higher SSD risk within AA-homozygotes.

These novel results highlight the endocannabinoid system’s role in the development and variability of dermatoglyphic patterns. The identified interaction encourages combining genetic and dermatoglyphics to assess neurodevelopmental alterations predisposing to SSD.

## 1. INTRODUCTION

Schizophrenia spectrum disorders (SSD) encompass different severe mental disorders, including schizophrenia, schizoaffective and schizophreniform disorders. The central etiological hypothesis sustains that SSD emerge as the consequence of multiple genetic and environmental factors altering the homeostasis of neurodevelopmental trajectories during the intrauterine and early postnatal periods, as well as during childhood and early adolescence (Kahn et al., 2015; Birnbaum and Weinberger, 2017).

During the early intrauterine neurodevelopmental process, the development of other ectodermal tissue derivates, such as dermatoglyphics, also occurs (Okajima, 1975; Babler, 1991; Fatjó-Vilas et al., 2008). The dermatoglyphic patterns are grooved configurations on palms and soles’ surface conformed by the alternation of epidermal ridges and sulci. These patterns are established from the 6^th^ to the 24^th^ week of gestation when their formation is complete, and they remain unchanged over the lifetime (Okajima, 1975; Babler, 1991). This process occurs in parallel with several crucial central nervous system development processes such as neural proliferation, cortex migration and prosencephalic development (Rakic, 1988; Volpe, 2000; Kalmady et al., 2015), so dermatoglyphics represent evidence of a particular neurodevelopmental window and dermatoglyphic alterations may be informative about early alterations in the neurodevelopmental process. The consideration of dermatoglyphics as indirect markers of neurodevelopmental alterations is supported by the high occurrence of dermatoglyphic deviations in chromosomal syndromes and neurodevelopment-related disorders caused by genetic and environmental factors (Schaumann Blanka and Alter, 1976). Considering the neurodevelopmental roots of SSD, several studies have reported quantitative and qualitative dermatoglyphic differences between patients affected by these disorders and healthy controls. Generally, patients tend to present simplified dermatoglyphic configurations and higher bilateral asymmetry (Fearon et al. 2001; Fañanas et al., 1996; Bramon et al., 2005). Indeed, dermatoglyphic pattern deviances have been highlighted as a relevant schizophrenia risk factor through an umbrella review (Radua et al., 2018).

Family and twin-based studies have determined dermatoglyphics heritability in variable but significant levels (h^2^=0.65 to 0.96) (Schaumann Blanka and Alter, 1976; Machado et al., 2010; Karmakar et al., 2011). While little is known about the dermatoglyphics-specific genetic determinants, a recent GWAS identified 18 loci associated with fingerprint type that highlighted the role of pathways related to limp development (Li et al., 2022). Nonetheless, information on to which extent the dermatoglyphics’ genetic and environmental determinants are shared with those of SSD is still missing. Among the mechanisms proposed to mediate gene and environment interactions on SSD and dermatoglyphic morphology, prenatal stress and obstetric complications achieve importance (Cannon et al., 2002; Byrne et al., 2007; Mittal et al., 2008). In this sense, several studies have shown a link between obstetric complications and dermatoglyphic ridge count reductions in schizophrenia (Bramon et al., 2005; Fatjó-Vilas et al., 2008). Then, foetal hypoxia seems to be a common denominator in the obstetric adverse events associated with psychosis based on gene-environment interaction studies and meta-analyses (Mittal et al., 2008; Davies et al., 2020).

Oxygen level variation is required for several physiological processes to take place, such as the formation of the neural fold (Cejudo-Martin and Johnson, 2005), the neural tube closure (Li et al., 2005) and oligodendrocyte proliferation and myelination (Barateiro et al., 2016). Accordingly, the regulation of hypoxia and its molecular mechanisms are considered essential for neurodevelopment and dysfunctions in such homeostasis may lead to abnormal gene expression and lasting changes in neuronal circuitry in the developing brain (Kietzmann et al., 2001). Interestingly, several reviews highlighted that among schizophrenia’s candidate genes, more than half of them met the criteria for a link to ischemia-hypoxia and/or vascular factors (Schmidt-Kastner et al., 2006, 2012).

Among the genes highlighted in Schmidt-Kastner’s studies (Schmidt-Kastner et al., 2006, 2012), there is the *CNR1* gene. An increasing body of evidence has emphasised the role of *CNR1* in the genetic underpinnings associated with schizophrenia (Gouvêa et al., 2017). It has also indicated the *CNR1* mediation between the influence of environmental risk factors and changes in brain structure and cognition in schizophrenia (Ho et al., 2011). The *CNR1* gene encodes for the CB1 receptor, the main endocannabinoid receptor in the brain and an essential central nervous system presynaptic receptor (Herkenham et al., 1990; Melis et al., 2004; Eggan and Lewis, 2007) with key effects on dopaminergic transmission (Fernández-Ruiz et al., 2010). Even though the role of the endocannabinoid system in epidermal differentiation has been barely studied, it is known that the CB1 receptor modulates human keratinocyte and epidermal differentiation, skin development (Maccarrone et al., 2003), and plays a role in the epidermal permeability barrier (Roelandt et al., 2012). Such particular functions of CB1 are integrated into the well-described roles of the endocannabinoid system in the regulation of cell-fate processes during development, including cell survival, proliferation, differentiation, and migration (Galve-Roperh et al., 2013; Gomes et al., 2020; Lu and Mackie, 2021). The CB1 receptors are expressed during early development in neuroepithelial progenitor cells (Galve-Roperh et al., 2013) and have been detected in the foetal brain as early as week 14 in regions of the frontal cortex, hippocampus, caudate nucleus, putamen, and cerebellum, mimicking their adult brain detection (Mato et al., 2003; Wang et al., 2003; Tao et al., 2020). In addition, the *CNR1* mRNA shows evidence of upregulation during ischemia (Jin et al., 2000), emphasising its expression sensitivity to oxygen levels.

Considering the shared genetic background between the development of the brain and dermatoglyphics and the role of the endocannabinoid system during these processes, we hypothesised that *CNR1* genetic variability influences both the dermatoglyphic configurations and the liability towards SSD and that it also modulates the relationship between dermatoglyphic pattern deviances and SSD risk. Then, we aimed to investigate the common underpinnings of SSD and dermatoglyphic patterns and combine phenomics of the dermatoglyphic patterns with the CNR1 genetic data to identify specific biomarkers for characterising the liability towards SSD.

## 2. METHODS

### 2.1. Sample

The sample consisted of 112 healthy controls (HC) (41(36.6%) males, mean age(sd)=25.85(6.54)) and 97 individuals diagnosed with a schizophrenia-spectrum disorder (SSD) (82.5% schizophrenia, 15.5% schizoaffective disorder and 2.1% schizophreniform disorder) (69 (71.1%) males, mean age(sd)=29.76(7.75)) according to DSM-IV-TR. All participants were of European ancestry. The exclusion criteria included chromosomal syndrome and, additionally, for HC, personal or family history of psychiatric service contact or treatment. Between cases and controls, there were differences regarding sex distribution (χ^2^=24.856, p<0.001) and age (U=3624.5 p<0.001).

Ethical approval was obtained from the Sainte Anne Hospital Research Ethics Committee, and all participants provided written informed consent about the study procedures and implications, which were carried out according to the Declaration of Helsinki.

### 2.2. Genotyping

All the individuals were genotyped for two SNPs in the *CNR1* gene (6q15): the rs2023239-A/G and the rs806379-A/T. These SNPs were selected based on their minor allele frequency in European Population (>5%), their potential relevance in relation to protein availability and their role in the pathophysiological mechanisms associated with SSD (Ketcherside et al. 2017, Zhang et al. 2005). Genomic DNA was extracted from buccal mucosa or peripheral blood cells using standard methods, and the selected variants were genotyped using TaqMan. The two variants were in Hardy-Weinberg equilibrium within both diagnostic groups (PLINK 1.07, (Purcell et al., 2007). The minor alleles in our sample matched the ones identified in the European Population from the 1000 Genomes, and their frequency was 0.35 for the rs2023239-G-allele and 0.35 for the rs806379-T-allele.

### 2.3. Dermatoglyphics assessment

Bilateral finger and handprints were obtained from all participants using specific methods (Speed Ball Block Ink, Utrecht Art Supplies, New Jersey, US). On the palms, we analysed the a-b ridge count (ABRC) of both hands, which is the number of ridges between the triradius *a* (in the base of the index finger) and the triradius *b* (in the base of the medium finder). A triradius is a Y-shaped point of convergence of ridges from 3 different directions. In the hand palm, triradii usually are in the base of the fingers 2 through 5. Then, we computed: i) the total a-b ridge count (TABRC, which is the addition of the right and the left ABRC) (**Figure 1.A**); and ii) the a-b ridge count fluctuating asymmetry (ABRC-FA), a measure of developmental instability, which is the absolute difference between the right and the left ABRC. On each finger, we identified the fingertip pattern: arches (associated with no triradius), loops (associated with one triradius) or whorls/double-loops (associated with two triradii). Then, we calculated the pattern intensity index (PII), which quantifies the number of triradii in the ten fingers and is a measure of the complexity of the finger configurations (Schaumann Blanka and Alter, 1976) (**Figure 1.B**).

**Figure 1.**
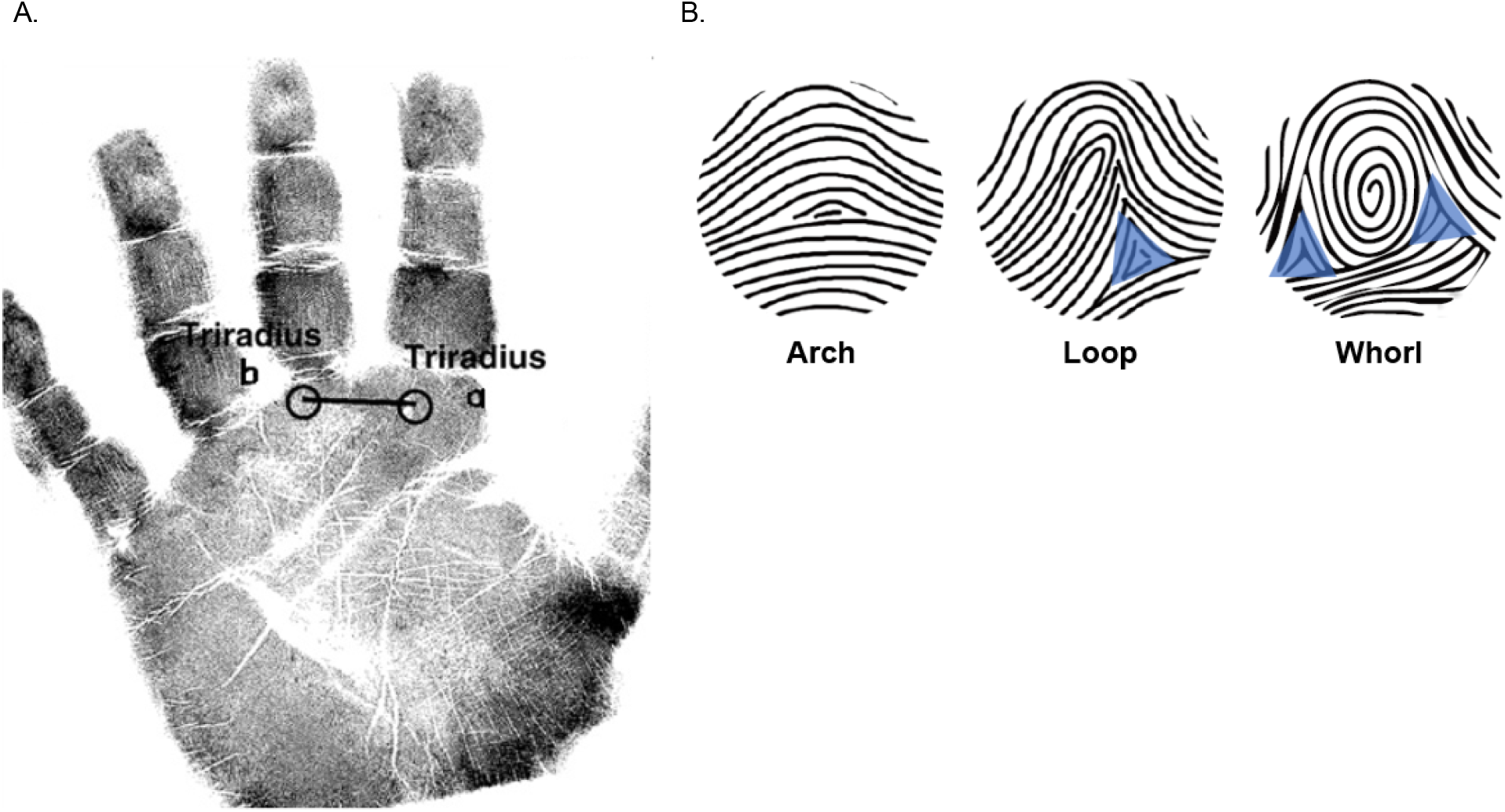
**A**. Handprints where the triradii a and b are indicated. The total a-b ridge count (TABRC) corresponds to the sum of the number of ridges between both triradii from the right and left hands. **B**. Different fingertip patterns (left to right): arch, loop and whorl with the triradius marked with blue triangles. Figures adapted from Fatjó-Vilas et al. 2008 and Li et al. 2022.

The final number of individuals analysed for each dermatoglyphic variable varies depending on the quality of the fingerprints. Dermatoglyphic variables assessment was performed by one of the authors (MFV), according to (Cummins and Midlo, 1943), blindly to individual diagnostic status. There were no dermatoglyphic differences between males and females or between groups (**Table 1**).

**Table 1.**
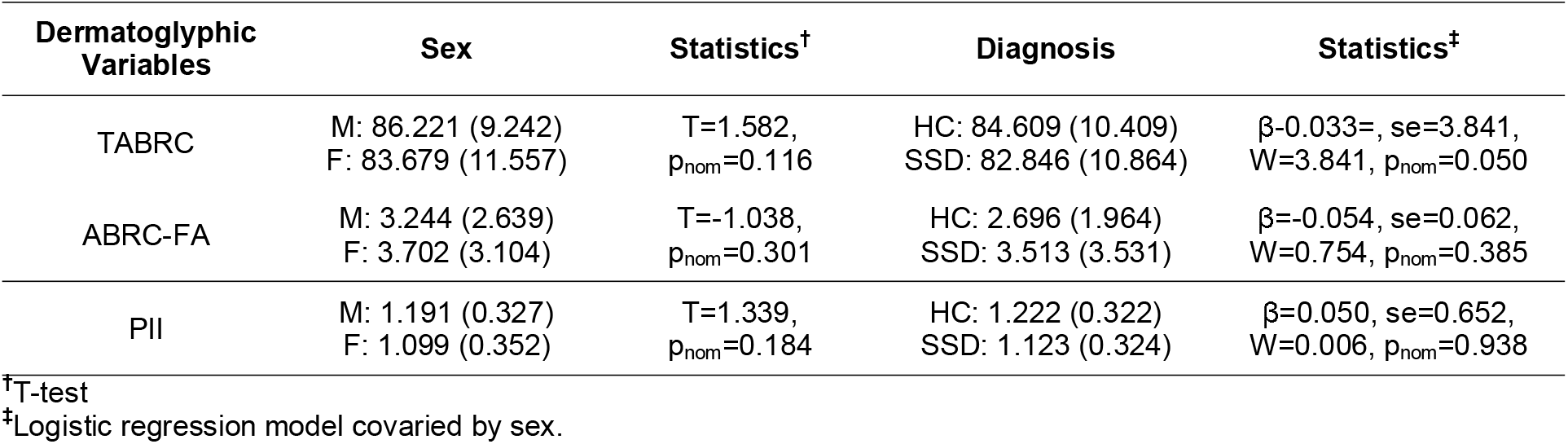
Description of the dermatoglyphic variability between males (M) and females (F), and between healthy controls (HC) and patients with schizophrenia-spectrum disorder (SSD). Mean and standard deviation (sd) are reported for the total a-b ridge count (TABRC), the a-b ridge count fluctuating asymmetry (ABRC-FA), and the pattern intensity index (PII) in both diagnostic groups.

### 2.4. Statistical analyses

All the data was processed in SPSS (SPSS 27.0, IBM SPSS Statistics for Windows, version 27.0, released 2020, IBM Corporation, Armonk, NY), and the genetic models were tested using PLINK. Tests for sex distribution and age differences across diagnostic categories were conducted using chi-square (χ^2^) and Mann-Whitney U (U) tests, respectively (SPSS).

Based on the sample distribution and to maximise the power, all the analyses were conducted assuming a minor allele dominance model. Then, the genotypes were dichotomised by grouping the minor and the heterozygous genotypes (rs2023230-AG/GG (G-allele carriers (Gcar)) *vs* rs2023239-AA, and rs806379-AT/TT (T-allele carriers (Tcar)) *vs* rs806379-TT).

Firstly, we examined the genetic association of *CNR1*-rs2023239 and *CNR1*-rs806379 genotypes with the risk for SSD. Secondly, we evaluated the effect of the genotypes on each dermatoglyphic measure separately in each group. Lastly, we explored whether there was a modulation effect of the *CNR1* variants on the relationship between dermatoglyphic variables and SSD vulnerability. For this purpose, we tested the interaction between the genotypes and each of the dermatoglyphic measures on the risk for SSD. To comprehend the effect of the significant interactions detected with PLINK, we subsequently obtained the corresponding predicted probabilities and plotted them (SPSS). All the analyses were conducted with logistic or linear regressions, when appropriate, and included sex as a covariate. Both the nominal (p_nom_) and the empirical (p_emp_) p-values, obtained after a 10,000 permutations procedure, are reported, and the significance threshold was set at p_emp_≤0.05.

## 3. RESULTS

Genetic association analysis on SSD showed that both *CNR1* polymorphisms were significantly associated with the disorders’ liability. Regarding rs2023239, we detected an overrepresentation of Gcar individuals among HC compared to patients; therefore, the AG/GG genotypes were associated with a protective effect. Concerning rs806379, we detected an overrepresentation of Tcar individuals among patients with SSD and the AT/TT genotypes were associated with a risk effect. The association results are displayed in **Table 2**.

**Table 2.** Significant results of the genotypic association analysis towards the risk for SSD (112 healthy controls (HC) and 97 patients with schizophrenia-spectrum disorders (SSD). The genotype frequencies are given for both diagnostic groups as well as the logistic regression statistics (Wald (W), OR and confidence interval [95%CI], and p-value after permutation).

| Genotypes | HC | Patients with SSD | Statistics | Significance |
| --- | --- | --- | --- | --- |
| rs2023239 (AA vs Gcar) | 0.30:0.70 | 0.59:0.41 | W=-3.995, OR<br>[95%CI]=0.285, [0.154:0.528] | $p_{nom}=6.46e-05$<br>$p_{emp}=2.00e-04$ |
| rs806379 (AA vs Tcar) | 0.57:0.43 | 0.29:0.71 | W=3.768, OR<br>[95%CI]=3.249, [1.760:5.996] | $p_{nom}=1.65e-4$<br>$p_{emp}=4.00e-04$ |

Subsequently, we inspected the genotypic effect on TABRC, ABRC-FA and PII within each diagnostic group. The data revealed effects within HC. First, the rs2023239-Gcar presented higher PII scores as compared to AA-homozygous (β=0.354, se=0.140, 95%CI=0.081:0.628, W=2.537, p_nom_=0.017, p_emp_=0.016, PII mean(sd) for Gcar=1.129(0.275) and AA=1.034(0.412)). Second, the rs806379-Tcar had lower TABRC as compared to AA-homozygous (β=-4.370, se=2.089, 95%CI=-8.464:-0.276, W=-2.092, p_nom_=0.039, p_emp_=0.038, TABRC mean(sd) for Tcar=83.430(11.754) and AA=87.490(9.529)). No *CNR1* effect on dermatoglyphic markers was detected in patients.

Lastly, we found that *CNR1*-rs2023239 variability significantly modulated the relationship between the PII and SSD risk. The full factorial model (including the rs2023239, the PII and their interaction) was globally significant (W=9.822, p_nom_=0.044, p_emp_=0.044), as well as the interaction term (OR=0.023, se=1.604, W=-2.361, OR[95%CI]=0.001:0.526, p_nom_=0.018, p_emp_=0.039). Subsequently, the predicted probabilities were obtained and plotted (**Figure 2**). The scatter plot showed that the relationship between the SSD risk and the PII was inverse depending on the rs2023239 genotype. More specifically, the individuals who were rs2023239-Gcar and had a higher PII presented a lower predicted probability towards SSD risk, while AA-homozygous and the same high levels of PII depicted the opposite relationship with the risk.

**Figure 2.**
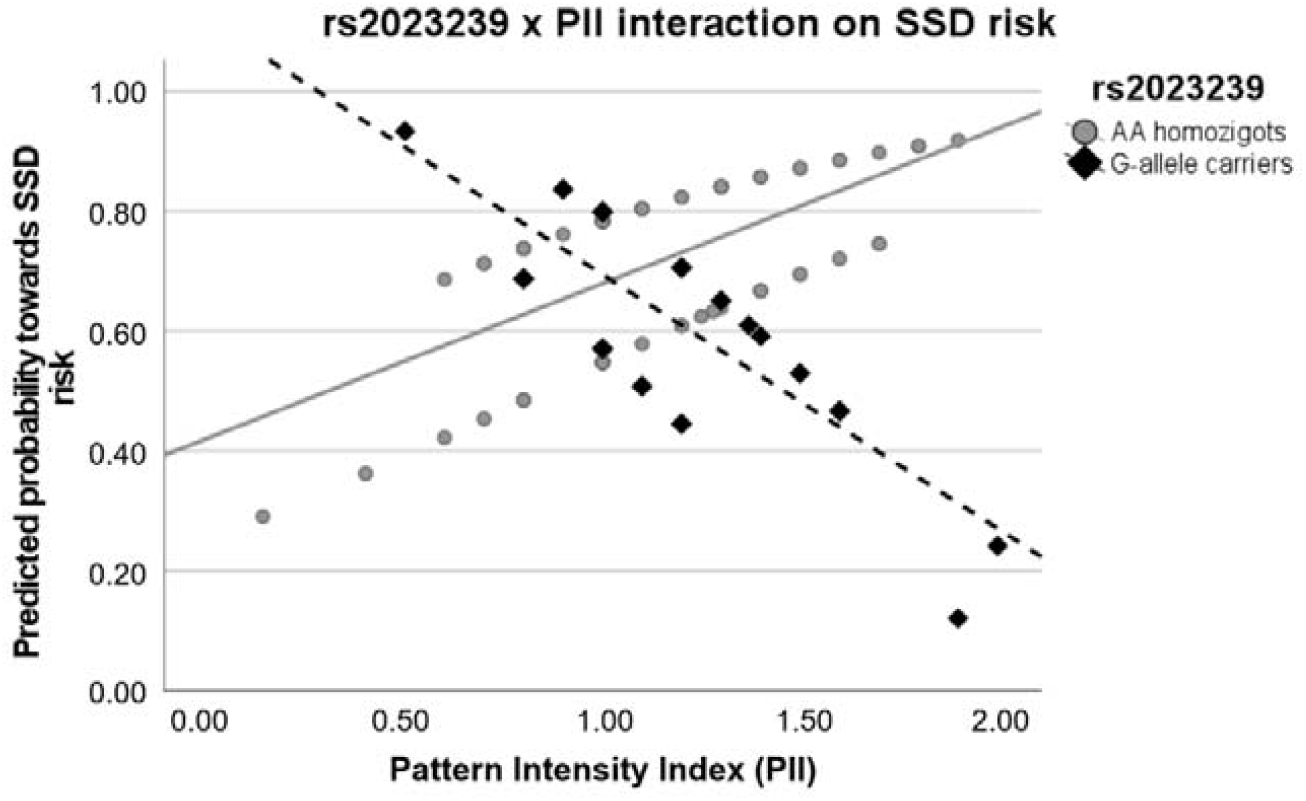
Interaction graph showing the interplay between the *CNR1*-rs2023239 genotype and the pattern intensity index (PII) on the risk for schizophrenia-spectrum disorders (SSD). A lower predicted probability indicates a lower risk towards SSD. Values show the inverse relationship between PII and predicted probability for SSD depending on the rs2023239 genotype.

## 4. DISCUSSION

This study aimed to investigate the shared genetic underpinnings of SSD and dermatoglyphic patterns by assessing whether the *CNR1* genetic variability influences the relationship between dermatoglyphic pattern deviances and SSD liability. To the best of our knowledge, this is the first study to explore the role of the endocannabinoid system in dermatoglyphic patterns variability, and the derived results point towards the combined effect of *CNR1* and dermatoglyphics on modulating the risk towards SSD.

First, our results evidence an influence of the two *CNR1* variants (rs2023239 and rs806379) on the predisposition towards SSD. Whereas many association studies have assessed the role of the *CNR1* variability on schizophrenia, SSD and other related clinical outcomes, existing data is very heterogeneous in terms of the population origin and the SNP selection (Gouvêa et al., 2017). Focusing on rs2023239, while there is no previous evidence of its association with the risk for psychosis *per se*, a modulation effect in the evolution of the psychopathological features and brain structural changes along the course of first-episode psychosis has been described (Suárez-Pinilla et al., 2015). Also, this polymorphism has been associated with the risk of metabolic syndrome in patients with SSD (Yu et al., 2013). On the other hand, previous studies failed to associate the rs806379 with the risk for schizophrenia in a Brazilian sample (Gouvêa et al., 2017), or with the risk for metabolic syndrome (Yu et al., 2013). However, considering other phenotypes associated with psychosis, evidence indicates that under exposure to early psychosocial adversity, rs806379 modulates impulsivity control in healthy adolescents (Buchmann et al., 2015). These associations may be driven by mechanisms unrelated to protein sequence since they lie in intronic regions but by expression regulatory mechanisms. In this sense, the rs2023239 seems to influence CB1 receptor density in lymphocytes (Ketcherside et al., 2017). If lymphocyte CB1 levels mimic the central nervous system, receptor availability changes could result in neurotransmission effects. Indeed, these two SNPs (rs2023239 and rs806379) in combination with another (rs1535255) have been associated with low levels of CB1 receptor mRNA in the cerebral cortex and the midbrain (Zhang et al., 2004), reinforcing the evidence of the modulatory effects of intronic variants on the receptor availability in the brain. It is also of interest that CB1 levels influence the expression of differentiation signals in various neuronal lineages (Galve-Roperh et al., 2013) and that several studies report altered endocannabinoid receptor concentrations in patients with schizophrenia in the dorsolateral prefrontal cortex, the posterior and anterior cingulate cortex (Dean et al. 2001; Zavitsanou et al., 2004; Newell et al., 2006; Tao et al., 2020).

Second, our findings link the *CNR1* rs2023239 and rs806379 variants with PII and TABRC variability in healthy controls. On the one hand, we describe that the rs2023239-G allele is associated with higher PII, which means that this allele, observed in less frequency in patients than in controls in our sample, is in turn related to higher dermatoglyphic complexity as represented by the presence of whorls and loops patterns, figures with more triradii. These results align with previous data reporting lower finger dermatoglyphic patterns complexity assessed through the frequency of the fingertip figures in schizophrenia and schizotypal traits (Chok et al., 2005; Arunpongpaisal et al., 2011; Norovsambuu et al., 2021). On the other hand, studies assessing TABRC as a developmental biomarker evidenced ridge count reductions in patients with schizophrenia and with SSD, as well as in subgroups of patients with reported perinatal complications (Fearon et al. 2001; Fañanas et al., 1996; Bramon et al., 2005; Fatjó-Vilas et al., 2008). Hence, the detected effect in controls would also agree with previous case-control association results.

We would have expected also to find a *CNR1* modulation effect on patients’ dermatoglyphic measures. Nonetheless, these results could reflect, on the one hand, the low expected penetrance that two single common variants have on these complex phenotypic measures. On the other, considering a multifactorial and polygenic context, we must consider the effect of different genetic and environmental forces underlying the dermatoglyphic configurations, as in the risk for psychosis (Bramon et al., 2005; Fatjó-Vilas et al., 2008; Van Os et al., 2008; Walder et al., 2014). Therefore, by analysing two SNPs, we focus on a particular biological pathway to assess its effect on patients’ dermatoglyphic complexity. Still, we should not forget that the global genetic makeup of the individual, together with the environmental context, shapes the developmental stability patterns (Bramon et al., 2005; Fatjó-Vilas et al., 2008). Then, our results could indicate, based on our findings in HC and not in patients, group differences in the sensitivity to gene-environmental insults along neurodevelopment processes and, particularly, in the effects that such group-specific ontogenetical patterns may have on dermatoglyphic markers.

Finally, we assessed whether *CNR1* modulated the dermatoglyphics SSD association. The analyses revealed an interplay between rs2023239 and PII on the liability for these disorders. Individuals who were Gcar and had higher PII presented a reduced liability for SSD contrarily to AA-homozygous with the same PII. The interaction results would again align with the case-control association data and the dermatoglyphic modulation effect found within HC. The findings highlight the role of the *CNR1* gene and the endocannabinoid system during neurodevelopment, mediating the environmental insults occurring along this process, and the data opens new venues for investigation. Further research analysing *CNR1* in depth would allow a better characterisation of this gene and its relationship with dermatoglyphic pattern variability and other neurodevelopmental biomarkers.

Our results relate the *CNR1* gene to a particular neurodevelopmental marker, the dermatoglyphic configurations. Many *CNR1* x environmental interactions have been described involving cannabis use, negative life events and childhood adversity on SSD susceptibility, SSD brain-based phenotypes and other mental disorders (Juhasz et al., 2009; Buchmann et al., 2015; Misiak et al., 2018). This, together with evidence suggesting a hypoxia modulation effect on *CNR1* mRNA levels (Jin et al., 2000), could lead to thinking about an interplay between *CNR1* and adverse prenatal environmental factors impacting the developmental trajectories reflected in dermatoglyphic and brain alterations. In this sense, future studies extending our data by assessing obstetric and perinatal complications would be of great value to evaluate the environmental influences on the brain and dermatoglyphic variables and pave the way for gene x dermatoglyphics studies in neurodevelopmental disorders.

Lastly, some limitations should be acknowledged. First, new analyses in larger samples are needed to confirm our findings. Despite the selection of two *CNR1* variants being made based on their relevance for SSD and neurodevelopment, the use of two SNPs neither represents the polygenic background of schizophrenia and SSD nor the genetic determinants of dermatoglyphic configurations. Further studies inspecting the role of genetic variants across the endocannabinoid system or even genome-wide on dermatoglyphic measurements captured through automated and multivariate approaches would help to comprehend the relationship between SSD and dermatoglyphics and to develop predictive statistical methodologies applied in the development of diagnostic tools.

In conclusion, our results add to previous evidence implicating *CNR1* with neurodevelopmental disorders such as SSD and represent new evidence on the role of this gene in the development and variability of dermatoglyphic patterns. These data align with the known role of the Cannabinoid receptor 1 in epidermal differentiation and skin development and its involvement in psychosis risk and environmental insults sensitivity. The results regarding the modulation role of *CNR1* in the dermatoglyphic markers and SSD risk relationship encourage new research on the combined use of genetic and dermatoglyphics for the assessment of neurodevelopmental alterations predisposing to SSD and may lead to the development of diagnostic instruments for the identification of subgroups of patients with a greater burden of neurodevelopmental alterations.

## Data Availability

All data produced in the present study are available upon reasonable request to the corresponding authors.

## ACKNOWLEDGEMENTS

This study received funding provided by: i) the Spanish Ministry of Science and Innovation, Instituto de Salud Carlos III (co-funded by the European Regional Development Fund (ERDF)/European Social Fund “Investing in your future”) through the project PI20/01002 to MF-V, through PFIS predoctoral contracts to MG-R (FI19/0352) and NH (FI21/00093) and, a Miguel Servet contract to MF-V (CP20/00072); ii) a PIF-Salut contract to AS-M (SLT017/20/000233), and; iii) the Comissionat per a Universitats i Recerca del DIUE of the Generalitat de Catalunya (Agència de Gestió d’Ajuts Universitats i de Recerca (AGAUR), 2021SGR1475).

## References

Arunpongpaisal S, Nanakorn Phd S, Mongconthawornchai Msc P, Virasiri S, Maneeganondh Bsc S, Thepsuthummarat Msc K. 2011. J Med Assoc Thai.

Babler WJ. 1991. Article in Birth Defects Original Article Series.

Barateiro A, Brites D, Fernandes A. 2016. Oligodendrocyte Development and Myelination in Neurodevelopment: Molecular Mechanisms in Health and Disease. Curr Pharm Des 22:656–679.

Birnbaum R, Weinberger DR. 2017. Genetic insights into the neurodevelopmental origins of schizophrenia. Nat Rev Neurosci 18:727–740.

Bramon E, Walshe M, McDonald C, Martín B, Toulopoulou T, Wickham H, Os J Van, Fearon P, Sham PC, Fañanás L, Murray RM. 2005. Dermatoglyphics and schizophrenia: A meta-analysis and investigation of the impact of obstetric complications upon a-b ridge count. Schizophr Res 75:399–404.

Buchmann AF, Hohm E, Witt SH, Blomeyer D, Jennen-Steinmetz C, Schmidt MH, Esser G, Banaschewski T, Brandeis D, Laucht M. 2015. Role of CNR1 polymorphisms in moderating the effects of psychosocial adversity on impulsivity in adolescents. J Neural Transm 122:455–463.

Byrne M, Agerbo E, Bennedsen B, Eaton WW, Mortensen PB. 2007. Obstetric conditions and risk of first admission with schizophrenia: A Danish national register based study. Schizophr Res 97:51–59.

Cannon M, Peter Jones MrcpB, Psych Robin Murray MM. 2002. Am J Psychiatry.

Cejudo-Martin P, Johnson RS. 2005. Developmental Cell.

Chok JT, Kwapil TR, Scheuermann A. 2005. Dermatoglyphic anomalies in psychometrically identified schizotypic young adults. Schizophr Res 72:205–214.

Cummins H, Midlo C. 1943. Finger Prints, Palms and Soles. An Introduction to Dermatoglyphics. The Blakiston Company, Philadelphia.

Davies C, Segre G, Estradé A, Radua J, Micheli A De, Provenzani U, Oliver D, Salazar de Pablo G, Ramella-Cravaro V, Besozzi M, Dazzan P, Miele M, et al. 2020. Prenatal and perinatal risk and protective factors for psychosis: a systematic review and meta-analysis. Lancet Psychiatry 7:399–410.

Dean B, Sundram S, Bradbury R, Scarr E, Copolov D. 2001. Studies on [3H]CP-55940 binding in the human central nervous system: regional specific changes in density of cannabinoid-1 receptors associated with schizophrenia and cannabis use. Neuroscience 103:9–15.

Eggan SM, Lewis DA. 2007. Immunocytochemical distribution of the cannabinoid CB1 receptor in the primate neocortex: A regional and laminar analysis. Cerebral Cortex 17:175–191.

Fañanas L, Os J Van, Hoyos C, Mcgrath J, Mellor CS, Murray R. 1996. Dermatoglyphic a-b ridge count as a possible marker for developmental disturbance in schizophrenia: replication in two samples. Schizophr Res 20:307–314.

Fatjó-Vilas M, Gourion D, Campanera S, Mouaffak F, Levy-Rueff M, Navarro ME, Chayet M, Miret S, Krebs MO, Fañanás L. 2008. New evidences of gene and environment interactions affecting prenatal neurodevelopment in schizophrenia-spectrum disorders: A family dermatoglyphic study. Schizophr Res 103:209–217.

Fearon P, Lane A, Airie M, Scannell J, Mcgowan A, Byrne M, Cannon M, Cotter D, Murphy P, Cassidy B, Waddington J, Larkin C, et al. 2001. Is reduced dermatoglyphic a±b ridge count a reliable marker of developmental impairment in schizophrenia? Schizophr Res 50:151–157.

Fernández-Ruiz J, Hernández M, Ramos JA. 2010. CNS Neuroscience and Therapeutics.

Galve-Roperh I, Chiurchiù V, Díaz-Alonso J, Bari M, Guzmán M, Maccarrone M. 2013. Progress in Lipid Research.

Gomes TM, Dias da Silva D, Carmo H, Carvalho F, Silva JP. 2020. Epigenetics and the endocannabinoid system signaling: An intricate interplay modulating neurodevelopment. Pharmacol Res 162:.

Gouvêa ES, Santos Filho AF, Ota VK, Mrad V, Gadelha A, Bressan RA, Cordeiro Q, Belangero SI. 2017. The role of the CNR1 gene in schizophrenia: A systematic review including unpublished data. Revista Brasileira de Psiquiatria 39:160–171.

Herkenham M, Lynn AB, Litrle MD, Johnsont MR, Melvin LS, Costa BR De, Riceo KC. 1990. Proc. Nati. Acad. Sci. USA.

Ho BC, Wassink TH, Ziebell S, Andreasen NC. 2011. Cannabinoid receptor 1 gene polymorphisms and marijuana misuse interactions on white matter and cognitive deficits in schizophrenia. Schizophr Res 128:66–75.

Jin KL, Mao XO, Goldsmith PC, Greenberg DA. 2000. CB1 cannabinoid receptor induction in experimental stroke. Ann Neurol 48:257–261.

Juhasz G, Chase D, Pegg E, Downey D, Toth ZG, Stones K, Platt H, Mekli K, Payton A, Elliott R, Anderson IM, Deakin JFW. 2009. CNR1 gene is associated with high neuroticism and low agreeableness and interacts with recent negative life events to predict current depressive symptoms. Neuropsychopharmacology 34:2019–2027.

Kahn RS, Sommer IE, Murray RM, Meyer-Lindenberg A, Weinberger DR, Cannon TD, O’Donovan M, Correll CU, Kane JM, Os J Van, Insel TR. 2015. Schizophrenia. Nat Rev Dis Primers 1:.

Kalmady S V., Shivakumar V, Gautham S, Arasappa R, Jose DA, Venkatasubramanian G, Gangadhar BN. 2015. Dermatoglyphic correlates of hippocampus volume: Evaluation of aberrant neurodevelopmental markers in antipsychotic-naïve schizophrenia. Psychiatry Res Neuroimaging 234:113–120.

Karmakar B, Malkin I, Kobyliansky E. 2011. Inheritance of 18 quantitative dermatoglyphic traits based on Factors in MZ and DZ twins. Anthropologischer Anzeiger 68:185–193.

Ketcherside A, Noble LJ, McIntyre CK, Filbey FM. 2017. Cannabinoid Receptor 1 Gene by Cannabis Use Interaction on CB1 Receptor Density. Cannabis Cannabinoid Res 2:202–209.

Kietzmann T, Knabe W, Schmidt-Kastner R. 2001. Hypoxia and hypoxia-inducible factor modulated gene expression in brain. Involvement in neuroprotection and cell death. Eur Arch Psychiatry Clin Neurosci 251:170–178.

Li J, Glover JD, Zhang H, Peng M, Tan J, Mallick CB, Hou D, Yang Y, Wu S, Liu Y, Peng Q, Zheng SC, et al. 2022. Limb development genes underlie variation in human fingerprint patterns. Cell 185:95–112.e18.

Li R, Chase M, Jung S-K, Smith PJS, Loeken MR. 2005. Hypoxic stress in diabetic pregnancy contributes to impaired embryo gene expression and defective development by inducing oxidative stress. Am J Physiol Endo-crinol Metab 289:591–599.

Lu HC, Mackie K. 2021. Biological Psychiatry: Cognitive Neuroscience and Neuroimaging.

Maccarrone M, Rienzo M Di, Battista N, Gasperi V, Guerrieri P, Rossi A, Finazzi-Agrò A. 2003. The endocannabinoid system in human keratinocytes: Evidence that anandamide inhibits epidermal differentiation through CB1 receptor-dependent inhibition of protein kinase C, activating protein-1, and transglutaminase. Journal of Biological Chemistry 278:33896–33903.

Machado JF, Fernandes PR, Roquetti RW, Fernandes Filho J. 2010. Digital dermatoglyphic heritability differences as evidenced by a female twin study. Twin Research and Human Genetics 13:482–489.

Mato S, Olmo E Del, Pazos A. 2003. Ontogenetic development of cannabinoid receptor expression and signal transduction functionality in the human brain. European Journal of Neuroscience 17:1747–1754.

Melis M, Pistis M, Perra S, Muntoni AL, Pillolla G, Gessa GL. 2004. Endocannabinoids Mediate Presynaptic Inhibition of Glutamatergic Transmission in Rat Ventral Tegmental Area Dopamine Neurons through Activation of CB1 Receptors. Journal of Neuroscience 24:53–62.

Misiak B, Stramecki F, Gawęda Ł, Prochwicz K, Sąsiadek MM, Moustafa AA, Frydecka D. 2018. Molecular Neurobiology.

Mittal VA, Ellman LM, Cannon TD. 2008. Schizophrenia Bulletin.

Newell KA, Deng C, Huang XF. 2006. Increased cannabinoid receptor density in the posterior cingulate cortex in schizophrenia. Exp Brain Res 172:556–560.

Norovsambuu O, Tsend-Ayush A, Lkhagvasuren N, Jav S. 2021. Main characteristics of dermatoglypics associated with schizophrenia and its clinical subtypes. PLoS One 16:.

Okajima M. 1975. Development of dermal ridges in the fetus. J Med Genet 12:243–250.

Os J Van, Rutten BPF, Poulton R. 2008. Schizophrenia Bulletin.

Purcell S, Neale B, Todd-Brown K, Thomas L, Ferreira MAR, Bender D, Maller J, Sklar P, Bakker PIW De, Daly MJ, Sham PC. 2007. PLINK: A tool set for whole-genome association and population-based linkage analyses. Am J Hum Genet 81:559–575.

Radua J, Ramella-Cravaro V, Ioannidis JPA, Reichenberg A, Phiphopthatsanee N, Amir T, Yenn Thoo H, Oliver D, Davies C, Morgan C, McGuire P, Murray RM, et al. 2018. World Psychiatry.

Rakic P. 1988. Specification of Cerebral Cortical Areas. Science (1979) 241:170–176.

Roelandt T, Heughebaert C, Bredif S, Giddelo C, Baudouin C, Msika P, Roseeuw D, Uchida Y, Elias PM, Hachem JP. 2012. Cannabinoid receptors 1 and 2 oppositely regulate epidermal permeability barrier status and differentiation. Exp Dermatol 21:688–693.

Schaumann Blanka and Alter M. 1976. Embryogenesis and Genetics of Epidermal Ridges. Dermatoglyphics in Medical Disorders, Berlin, Heidelberg: Springer Berlin Heidelberg, p 1–11.

Schmidt-Kastner R, Os J Van, Esquivel G, Steinbusch HWM, Rutten BPF. 2012. Molecular Psychiatry.

Schmidt-Kastner R, Os J van, W.M. Steinbusch H, Schmitz C. 2006. Schizophrenia Research.

Suárez-Pinilla P, Roiz-Santiañez R, Ortiz-García de la Foz V, Guest PC, Ayesa-Arriola R, Córdova-Palomera A, Tordesillas-Gutierrez D, Crespo-Facorro B. 2015. Brain structural and clinical changes after first episode psychosis: Focus on cannabinoid receptor 1 polymorphisms. Psychiatry Res Neuroimaging 233:112–119.

Tao R, Li C, Jaffe AE, Shin JH, Deep-Soboslay A, Yamin R, Weinberger DR, Hyde TM, Kleinman JE. 2020. Cannabinoid receptor CNR1 expression and DNA methylation in human prefrontal cortex, hippocampus and caudate in brain development and schizophrenia. Transl Psychiatry 10:.

Volpe J. 2000. Overview: normal and abnormal human brain development. Ment Retard Dev Disabil Res Rev 6:1–5.

Walder DJ, Faraone S V., Glatt SJ, Tsuang MT, Seidman LJ. 2014. Genetic liability, prenatal health, stress and family environment: Risk factors in the Harvard Adolescent Family High Risk for Schizophrenia Study. Schizophr Res 157:142–148.

Wang X, Dow-Edwards D, Keller E, Hurd YL. 2003. Preferential limbic expression of the cannabinoid receptor mRNA in the human fetal brain. Neuroscience 118:681–694.

Yu W, Hert M De, Moons T, Claes SJ, Correll CU, Winkel R Van. 2013. CNR1 gene and risk of the metabolic syndrome in patients with schizophrenia. J Clin Psychopharmacol 33:186–192.

Zavitsanou K, Garrick T, Huang XF. 2004. Selective antagonist [3H]SR141716A binding to cannabinoid CB1 receptors is increased in the anterior cingulate cortex in schizophrenia. Prog Neuropsychopharmacol Biol Psychiatry 28:355–360.

Zhang PW, Ishiguro H, Ohtsuki T, Hess J, Carillo F, Walther D, Onaivi ES, Arinami T, Uhl GR. 2004. Human cannabinoid receptor 1: 5′ exons, candidate regulatory regions, polymorphisms, haplotypes and association with polysubstance abuse.

